# No association between the *Plasmodium vivax crt-o* MS334 or In9*pvcrt* polymorphisms and chloroquine failure in a clinical cohort from Malaysia

**DOI:** 10.1101/2022.11.30.22282917

**Authors:** Angela Rumaseb, Roberto R. Moraes Barros, Juliana M. Sá, Jonathan J. Juliano, Timothy William, Kamil A. Braima, Bridget E. Barber, Nicholas M Anstey, Ric N. Price, Matthew J. Grigg, Sarah Auburn, Jutta Marfurt

## Abstract

Increasing reports of resistance to a frontline malaria blood-stage treatment, chloroquine (CQ), raise concerns for the elimination of *Plasmodium vivax*. The absence of an effective molecular marker of CQ resistance in *P. vivax* greatly constrains surveillance of this emerging threat. A recent genetic cross between CQ sensitive (CQS) and CQ resistant (CQR) NIH-1993 strains of *P. vivax* linked a moderate CQR phenotype with two candidate markers in *P. vivax* CQ resistance transporter gene (*pvcrt-o*): MS334 and In9*pvcrt*. Longer TGAAGH motifs at MS334 were associated with CQ resistance, as were shorter motifs at the In9*pvcrt* locus. In this study, high-grade CQR clinical isolates of *P. vivax* from Malaysia were used to investigate the association between the MS334 and In9*pvcrt* variants and treatment efficacy. Amongst a total of 49 independent monoclonal *P. vivax* isolates assessed, high-quality MS334 and In9*pvcrt* sequences could be derived from 30 (61%) and 23 (47%), respectively. Five MS334 and six In9*pvcrt* alleles were observed, with allele frequencies ranging from 2 to 76% and 3 to 71%, respectively. None of the clinical isolates had the same variant as the NIH-1993 CQR strain, and none were associated with CQ treatment failure (all *p*>0.05). Our findings suggest that the *pvcrt-o* MS334 and In9*pvcrt* markers cannot be used universally as markers of CQ treatment efficacy in an area of high-grade CQ resistance. Further studies applying hypothesis-free genome-wide approaches are warranted to identify more effective CQR markers for *P. vivax*.

## Introduction

*Plasmodium vivax* malaria remains an important public health burden affecting the poorest and most vulnerable communities of more than 49 endemic countries (1, 2). Chloroquine (CQ) is still the most widely used antimalarial drug to treat *P. vivax* but increasing reports of chloroquine resistance highlight the importance of surveillance of its clinical efficacy (3). Although surveillance of clinical efficacy is the gold standard for the detection of CQ resistance, challenges in discriminating infections deriving from blood stage failures (recrudescence), new mosquito inoculations (reinfections) and reactivation of dormant liver stages (relapses) confounds robust clinical estimates of CQ resistance (4). Molecular surveillance presents a more accurate and cost-effective strategy to identify drug resistant infections (5). However, the genetic basis of CQ resistance in *P. vivax* remains unclear.

Several studies have investigated Single Nucleotide Polymorphisms (SNPs) in orthologues of genes involved in CQ resistance in *P. falciparum*, namely *pvcrt-o* and *pvmdr1*, but none of these markers have been confirmed to be associated with clinical outcomes following CQ treatment of *P. vivax* (6-9). Other studies have reported an association between *pvcrt-o* expression levels and *P. vivax* CQ resistance (10-13). However, no evidence of association was observed between *pvcrt-o* expression and *ex vivo* drug susceptibility phenotypes using clinical isolates from Papua Indonesia (14), an area where high rates of clinical failure have been documented following CQ treatment (15).

More recently, a genetic cross between *P. vivax* subpopulations differing in susceptibility to CQ in non-human primates identified two new, non-coding *pvcrt-o* markers associated with *in vivo* CQ sensitivity: a repeat length polymorphism (MS334) and an intron 9 polymorphism (In9*pvcrt*) (12). The MS334 locus is located approximately 0.6 Kb upstream of the start codon of *pvcrt-o;* CQ sensitive (CQS) progeny contained 10 TGAAGH motifs, whilst CQ resistant (CQR) progeny carried a longer fragment of 15 TGAAGH motifs. At the In9*pvcrt* locus, 17 TGAAGH motifs were present in CQS progeny, whereas a shorter fragment comprising 14 TGAAGH motifs were present in CQR progeny.

The aim of the current study was to correlate the *pvcrt-o* MS334 and In9*pvcrt* markers with clinical outcomes following CQ treatment in patients enrolled into a prospective clinical trial conducted in Sabah, Malaysia (16). In this trial more than 60% of patients had recurrent parasitaemia within 28 days of treatment, indicative of high-grade CQ resistance (3, 4, 16).

## Materials and Methods

### Sample Collection and Selection

Clinical isolates of *P. vivax* were collected from a comparative efficacy study of patients with uncomplicated vivax malaria conducted in Sabah, Malaysia, between 2012 and 2014 (Clinical Trials Registration number NCT01708876) (16). Briefly, consenting patients aged ≥1 year and weighing >10kg, presenting with acute, uncomplicated vivax malaria to the study hospitals in Kudat, Kota Marudu and Pitas were recruited into the prospective study. Patients were randomly assigned to either CQ (median total dose of 27.7 mg/kg [range, 25.0–33.0]) or artesunate-mefloquine (AS-MQ) (median 11.0 mg/kg [range, 9.8–12.0] AS and 27.6 mg/kg [range, 24.6– 30.0] MQ), and followed for 42 days. Administration of primaquine for radical cure was delayed until 28 days of follow up. All treatment doses of chloroquine were supervised. The primary outcome was the cumulative risk of *P. vivax* parasitaemia by day 28. Blood samples were collected at enrolment and on the day of recurrence for molecular analyses. Monoclonal *P. vivax* isolates collected at enrolment into the CQ treatment were selected for further molecular analysis in the *pvcrt-o* study. Isolates were categorized as CQS if the patient exhibited adequate parasite clearance and no recurrent parasitaemia within 42 days. If patients failed to clear peripheral parasitaemia on blood smear or presented with recurrent *P. vivax* parasitaemia within 42 days after CQ treatment in the presence of a minimum effective plasma concentration of chloroquine and desethylchloroquine (CQ+DCQ) of ≥15ng/mL at Day 7 (corresponding to a whole blood concentration of 100ng/mL) (17), they were defined as CQR (16-18). All patient samples and corresponding clinical data were assigned unique study code identifiers (not identifiable to anyone outside the research group) to protect the identity of the subjects.

### DNA Extraction, Speciation, and Genotyping

DNA was extracted from 200 μL whole blood using the QIAamp DNA Blood extraction kit according to the manufacturer’s procedures (Qiagen, Germany). *Plasmodium spp*. was confirmed using the multiplex PCR method described by Padley et al. (19). *P. vivax* genotyping data was derived from published data stored in the open access vivaxGEN-MS platform (https://vivaxgen.menzies.edu.au) using batch code MYPV-MG and applying the default parameters for genotype calling (20). Briefly, genotyping was performed using capillary electrophoresis at nine short tandem repeat (STR) markers to determine the multiplicity of infection (MOI) as previously described (21).

### *pvcrt-o* Gene Fragment Amplification and Sanger Sequencing

The primer sequences and PCR cycling conditions are summarised in **Table 1**. The PCR products were run on 2% agarose gel, stained with SYBR™ Safe DNA Gel Stain (Invitrogen, USA) and visualized using the Molecular Imager Gel Doc XR system (Bio-Rad, USA). PCR amplification of In9*pvcrt* yielded multiple bands and henceforth gel extraction was required prior to sequencing, as conducted on previous experiments. Amplicons were sent to Macrogen Inc (Seoul, South Korea), where PCR product purification by gel extraction was conducted prior to sequencing using the dideoxy termination method and the respective PCR primers.

**Table 1.**
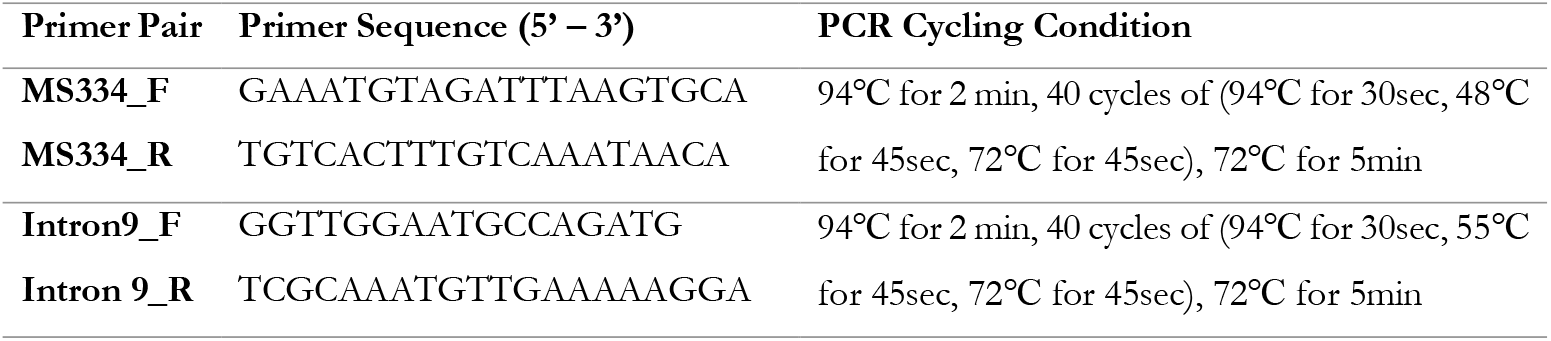
Primer sequences and PCR cycling conditions. PCR = Polymerase Chain Reaction

### Sequence Analysis

Trimming low-quality ends of the sequences were performed using Mega-X software (Version 10.2.5). Post-trimming, manual assembly of contigs and comparison with the gene regions of the CQS reference Sal-1 strain were performed using BioEdit software (Version 7.2.5). Alignments were undertaken with the bioinformatician blinded to the CQ treatment outcomes of the patients. To ensure consistency in assemblies between the genetic cross study (12) and the current study, the NIH-1993-S and NIH-1993-R sequences described in the former study were analysed alongside the Malaysian assemblies.

### Statistical Analysis

All statistical tests were conducted using in-built functions of R (Version 1.2.5042). Proportions were assessed using the Chi-squared test with Yates’ correction and Wilcoxon test was used for non-parametric comparisons. A significance threshold of *p<*0.05 was used for all statistical comparisons.

### Ethics

All samples were collected with written informed consent from patient or legal guardian for individuals less than 18 years of age. Ethical approval for the patient sampling and parasite molecular analysis was provided by the Human Research Ethics Committee of the Northern Territory Department of Health and Families (HREC-2010-1431, HREC-2012-1815 and HREC-2010-1396), the National Medical Research Ethics Committee, Ministry of Health, Malaysia (NMMR-10-754-6684, NMRR-12-511-12579).

## Results

### Summary of patient samples and sequencing data

The filtering process for all samples used in the study is presented in **Figure 1**. Amongst 49 clinical samples collected at baseline from patients treated with CQ, high-quality sequences could be derived from independent monoclonal *P. vivax* infections for MS334 in 30 (61%) isolates and In9*pvcr*t in 23 (47%) isolates. An additional 9 high-quality sequences at MS334 and 3 at In9*pvcrt* were also derived in paired isolates at baseline and day of recurrence. Details of the clinical characteristics of the patients and assay results of the isolates are presented in **Supplementary File 1**. There were no significant differences in age, gender, and baseline parasitaemia between the *pvcrt-o* study set and all patients treated with CQ in the trial (**Table 2**).

**Figure 1.**
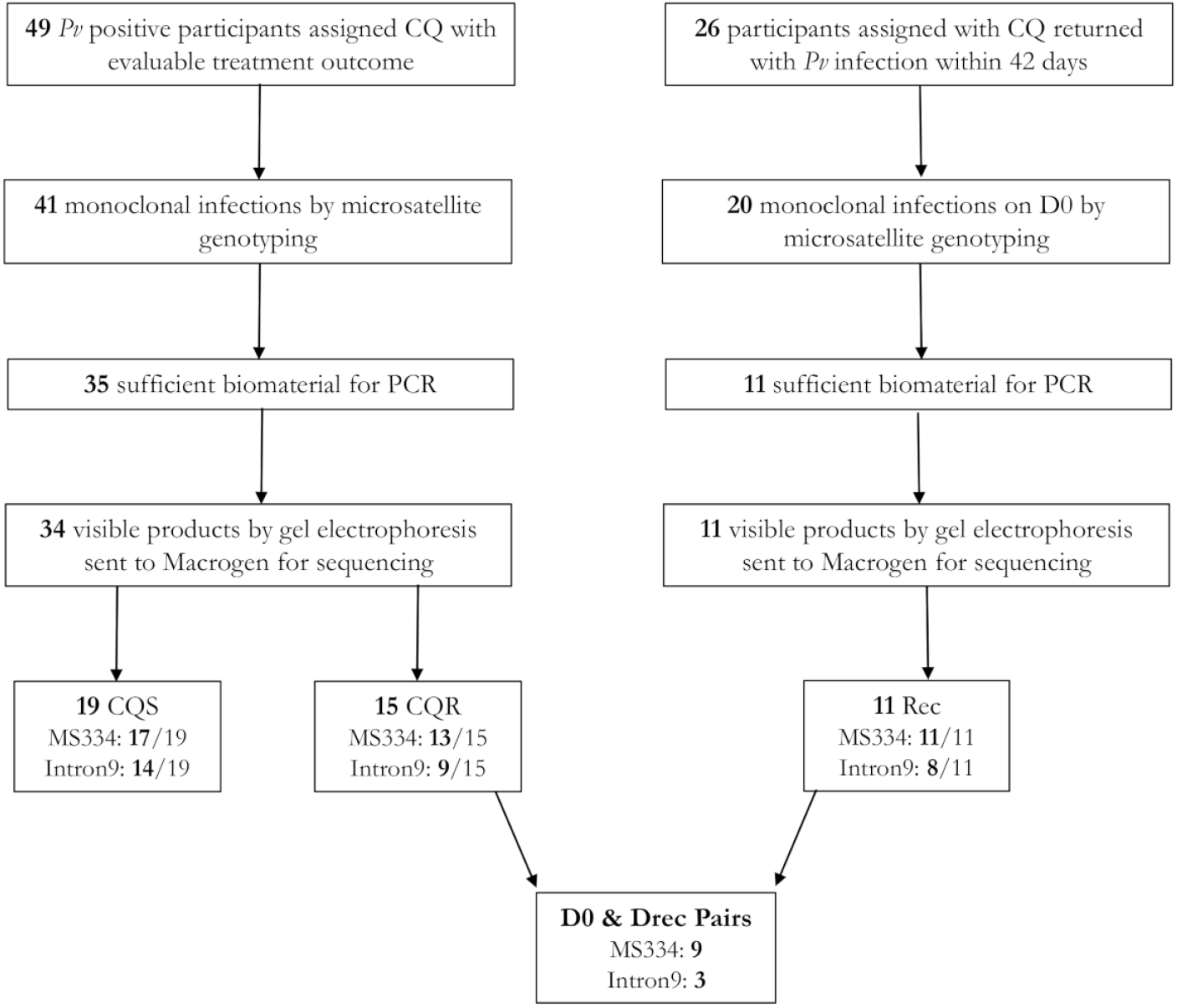
Flowchart outlining patient sample exclusion. Abbreviations: *Pv, Plasmodium vivax;* CQ, Chloroquine; PCR, polymerase chain reaction; CQS, chloroquine sensitive; CQR, chloroquine resistant; Rec, recurrent infection time point.

**Table 2.**
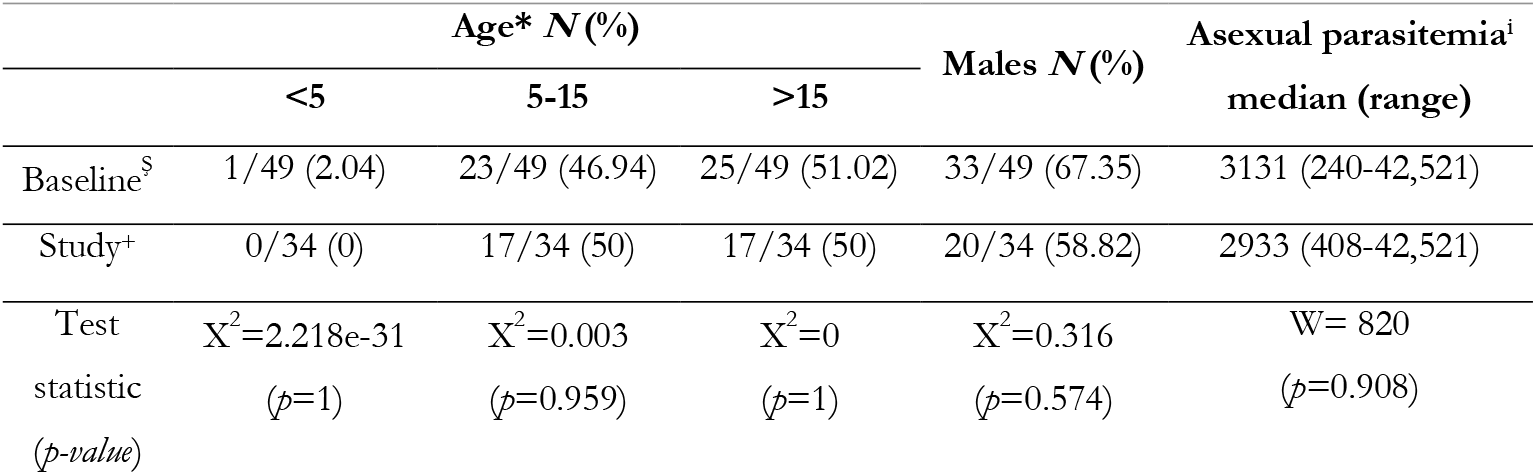
Demographic data of CQ-assigned participants in clinical trial and *pvcrt-o* study set. Symbols: *Age in years, ^i^Parasites/μL, ^Ş^ Participants assigned CQ on baseline, ^+^Participants included in the *pvcrt-o* study set.

|  | Age* N(%) |  |  | Males N(%) | Asexual parasitemia<sup>i</sup><br>median (range) |
| --- | --- | --- | --- | --- | --- |
|  | <5 | 5-15 | >15 |  |  |
| Baseline<sup>s</sup> | 1/49 (2.04) | 23/49 (46.94) | 25/49 (51.02) | 33/49 (67.35) | 3131 (240-42,521) |
| Study<sup>+</sup> | 0/34 (0) | 17/34 (50) | 17/34 (50) | 20/34 (58.82) | 2933 (408-42,521) |
| Test | X<sup>2</sup>=2.218e-31 | X<sup>2</sup>=0.003 | X<sup>2</sup>=0 | X<sup>2</sup>=0.316 | W= 820 |
| statistic | (p=1) | (p=0.959) | (p=1) | (p=0.574) | (p=0.908) |
| (p-value) |  |  |  |  |  |

Sequencing data on the successfully sequenced and aligned MS334 and In9*pvcrt* amplicons are deposited in Genbank under accession numbers OP807877 - OP807946. The sequencing pass rates amongst the baseline and recurrent infections were 91% (41/45) for MS334 and 69% (31/45) for In9*pvcrt*.

### Summary of MS334 and In9*pvcrt* variants observed in Malaysia

A summary of the MS334 and In9*pvcrt* genotype calls is provided in **Table 3**. Five MS334 and six In9*pvcrt* insertion and deletion (indel) variants were identified amongst the Day 0 and recurrent Malaysian isolates, with allele frequencies ranging from 2% (1/41) to 76% (31/41) and 3% (1/31) to 71% (22/31), respectively. Representative sequences of the 5 MS334 and 6 In9*pvcrt* indels observed in Malaysia, as well as the NIH-1993-R and NIH-1993-S strains, aligned against the Sal-1 reference strain, are provided in **Supplementary Files 2 and 3**.

**Table 3.**
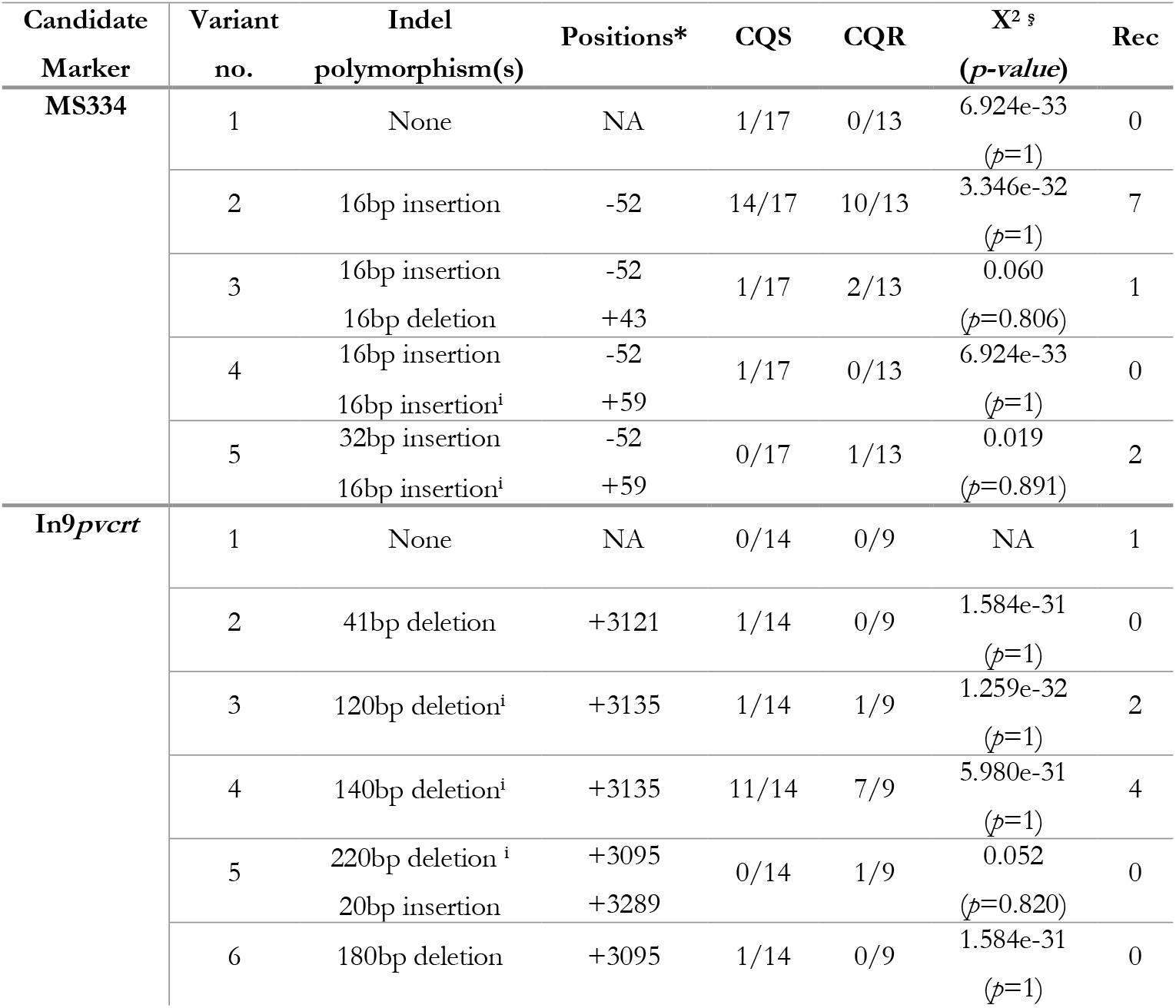
Summary of insertion and deletion variants observed in Malaysia. Abbreviations and symbols: Indels, Insertion and deletion; *Number of base pairs upstream (-) or downstream (+) from the start of *pvcrt-o* in Sal-1 strain (Pv_Sal1_chr01:330,260); ^**ş**^ Chi-squared test of CQS vs CQR baseline (i.e., not including the recurrent time point) isolates; ^i^ Part of polymorphisms observed in NIH-1993-R mentioned by (12); CQS, chloroquine sensitive; CQR, chloroquine resistant; Rec, recurrent infection time point. See **Supplementary Files 2 and 3** for sequence details for each variant.

At the MS334 locus, longer TGAAGH motifs were associated with CQR in the NIH-1993-R×S strains (12). The NIH-1993-R type (inferred CQR type) MS334 allele comprising the 15 TGAAGH motifs was not observed in Malaysia, and all variants at this locus exhibited shorter motif lengths, ranging from 8-11 motifs. Most clinical isolates also contained motifs shorter than the NIH-1993-S (10 motifs) type.

At the In9*pvcrt* locus, shorter TGAAGH motifs were associated with CQR in the NIH-1993 strains (12). The NIH-1993-R type In9*pvcrt* allele comprising 14 TGAAGH motifs was not observed in the clinical isolates from Malaysia, but motif lengths ranging from 7-17 were observed. Most infections contained 10 TGAAGH motifs (22/31; 71%), which is shorter than both the NIH-1993-R (14 motifs) and NIH-1993-S (17 motifs) types. The distribution of the different TGAAGH (TGAAGC, TGAAGA, TGAAGT) motif lengths observed in clinical CQS, CQR and recurrent isolates are summarised in **Figure 2**.

**Figure 2.**
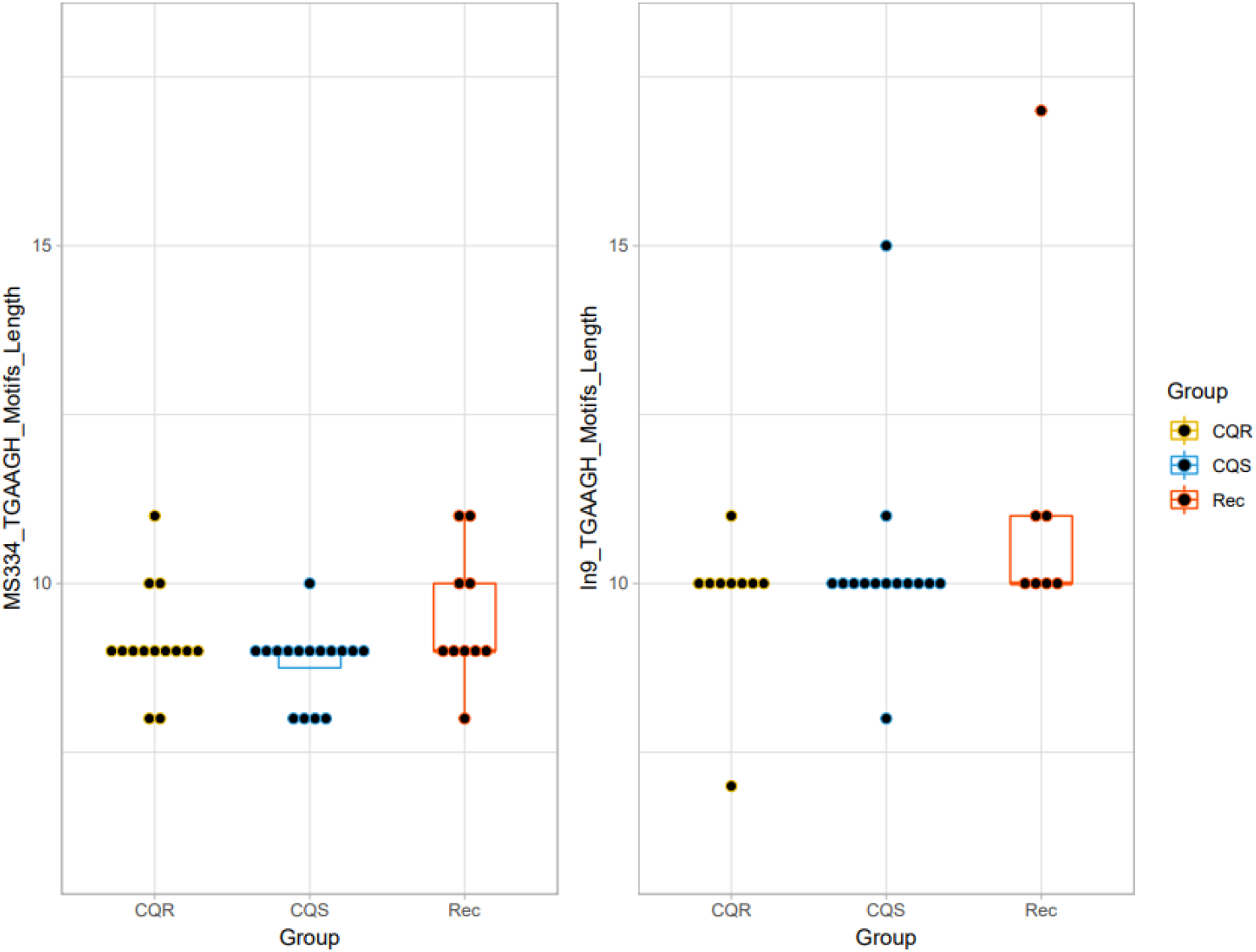
Dot-boxplots of TGAAGH motif lengths observed in Malaysian isolates. Abbreviations: CQS, chloroquine sensitive (day 0 infections); CQR, chloroquine resistant (day 0 infections); Rec, recurrent infection time point.

### Assessment of MS334 and In9*pvcrt* association with CQ treatment outcome

There was no statistically significant difference in the proportion of any of the five MS334 or six In9*pvcrt* alleles (all *p*>0.05) between CQS and CQR clinical isolates; **Table 3**. Although none of the clinical isolates carried the exact motifs of the NIH-1993-R strains at the MS334 and In9*pvcrt* loci, the Malaysian MS334 variant types 4 and 5, and In9*pvcrt* variant types 3, 4 and 5 possessed fragments of the NIH-1993-R indels (**Supplementary Files 2 and 3**). However, there was no significant difference in the proportion of the combined MS334 variants4 and 5 or combined In9*pvcrt* variants 3, 4 and 5 between the CQS and CQR isolates as summarised in **Table 4**.

**Table 4.**
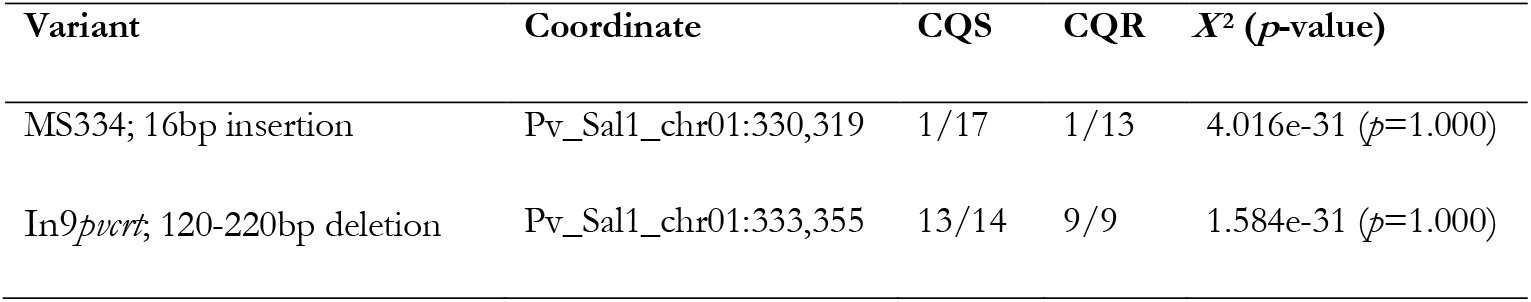
Summary of NIH-1993-R-type insertion and deletion polymorphisms observed in Malaysia. Abbreviations and symbols: CQS, chloroquine sensitive; CQR, chloroquine resistant.

Further assessment of the MS334 and In9*pvcrt* variants was undertaken in paired isolates at baseline and the day of recurrent parasitaemia. Seven of the 9 pairs successfully sequenced at MS334 and all 3 pairs sequenced at In9*pvcrt* exhibited the same alleles at both time points (**Table 5**). In the two pairs exhibiting allelic differences at MS334 (MV019 and TK026 pairs), the changes in variants pre- and post-treatment were not consistent. One pair possessed a longer allele at baseline, and the other pair possessed a longer allele at recurrence. Analysis of microsatellite-based genotyping data in two paired isolates revealed evidence of polyclonal infection at recurrence but not at baseline (**Table 6**).

**Table 5.**
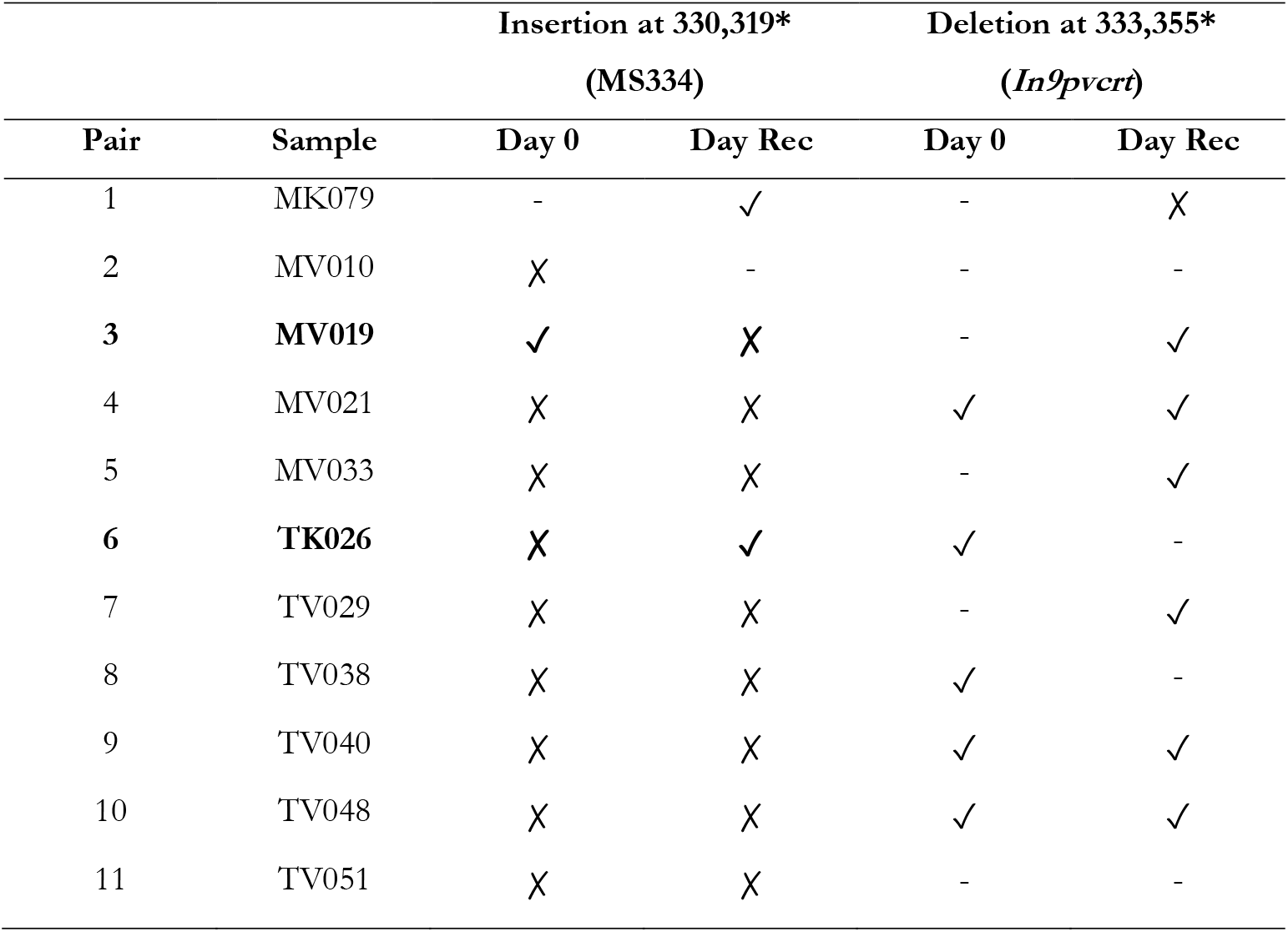
Summary of NIH-1993-R-type insertion and deletion polymorphisms observed in paired Malaysian isolates. Abbreviations and symbols: * Positions at chromosome 1 of Sal-1 reference strain; Rec, recurrent infection time point; (-) not amplified; (✓) insertion/deletion is present; (✗) insertion/deletion is not present. Infections with day 0 versus recurrence differences are highlighted **in bold**.

**Table 6.** Comparison of microsatellite genotyping data and *pvcrt-o* MS334 and In9*pvcrt* markers results in paired Malaysian isolates. Infections with day 0 versus recurrence differences are highlighted in bold. * Microsatellite-based multi-locus genotype (MLG).

| Pair | Sample | MS12 | pv3.27 | msp1f3 | MS10 | MS5 | MS1 | MS8 | MS16 | MS20 | MLG* | Indel Mutation Similarities |  |
| --- | --- | --- | --- | --- | --- | --- | --- | --- | --- | --- | --- | --- | --- |
|  |  |  |  |  |  |  |  |  |  |  |  | MS334 | *In9pvcrt* |
| 1 | MK079 | 209 | 280 | 258 | 195 | 194 | 237 | 281 | 385 | 201 | 1 | - | - |
| 1 | MK2079 | 209 | 280 | 258 | 195 | 194 | 237 | 281 | 385 | 201 | 1 | - | - |
| 2 | MV010 | 206 | 280 | 261 | 180 | 182 | 237 | 263 | 352 | 201 | 2 | - | - |
| 2 | MV1010 | 206 | 280 | 261 | 180 | 182 | 237 | 263 | 352 | 201 | 2 | - | - |
| **3** | **MV019** | **209** | **280** | **258** | **195** | **182** | **237** | **281** | **385** | **219** | **3** | **X** | - |
| **3** | **MV1019** | **209** | **280** | **258** | **195/153** | **182** | **237** | **281** | **385** | **219** | **3/4** | **X** | - |
| 4 | MV021 | 209 | 280 | 348 | 195 | 164 | 228 | 281 | 463 | 201 | 5 | ✓ | ✓ |
| 4 | MV1021 | 209 | 280 | 348 | 195 | 164 | 228 | 281 | 463 | 201 | 5 | ✓ | ✓ |
| 5 | MV033 | 209 | 280 | 255 | 195 | 173 | 237 | 281 | 418 | 201 | 6 | ✓ | - |
| 5 | MV1033 | 209 | 280 | 255 | 195 | 173 | 237 | 281 | 418 | 201 | 6 | ✓ | - |
| **6** | **TK026** | **209** | **280** | **255** | **195** | **173** | **237** | **281** | **418** | **201** | **6** | **X** | - |
| **6** | **TK1026** | **209** | **280** | **255** | **195** | **173/182** | **237** | **281** | **418** | **201** | **6/7** | **X** | - |
| 7 | TV029 | 209 | 280 | 255 | 195 | 173 | 237 | 281 | 418 | 201 | 6 | ✓ | - |
| 7 | TV1029 | 209 | 280 | 255 | 195 | 173 | 237 | 281 | 418 | 201 | 6 | ✓ | - |
| 8 | TV038 | 209 | 280 | 255 | 195 | 173 | 237 | 281 | 418 | 201 | 6 | ✓ | - |
| 8 | TV1038 | 209 | 280 | 255 | 195 | 173 | 237 | 281 | 418 | 201 | 6 | ✓ | - |
| 9 | TV040 | 209 | 280 | 255 | 195 | 173 | 237 | 281 | 418 | 201 | 6 | ✓ | ✓ |
| 9 | TV1040 | 209 | 280 | 255 | 195 | 173 | 237 | 281 | 418 | 201 | 6 | ✓ | ✓ |
| 10 | TV048 | 209 | 280 | 255 | 195 | 173 | 237 | 281 | 418 | 201 | 6 | ✓ | ✓ |
| 10 | TV1048 | 209 | 280 | 255 | 195 | 173 | 237 | 281 | 418 | 201 | 6 | ✓ | ✓ |
| 11 | TV051 | 209 | 280 | 255 | 195 | 173 | 237 | 281 | 418 | 201 | 6 | ✓ | - |
| 11 | TV1051 | 209 | 280 | 255 | 195 | 173 | 237 | 281 | 418 | 201 | 6 | ✓ | - |

## Discussion

The involvement of *pvcrt-o* in conferring CQ resistance in *P. vivax* has been investigated in several studies over the past two decades but remains contentious (8, 10-14, 22). A recent study using a genetic cross between CQS and moderate (surviving paediatric CQ doses) CQR parasite subpopulations of the NIH-1993-S×R strain linked *pvcrt-o* transcription to CQ resistance and two candidate markers in the gene region (MS334 and In9*pvcrt*)(12). Our study provides the first investigation of the MS334 and In9*pvcrt* variants using samples from a CQ clinical efficacy trial (16). Using patient isolates from a clinical trial of CQ efficacy on *P. vivax* undertaken in Sabah, Malaysia, a region with high-grade CQR during the sample collection period and at a time of residual vivax endemicity, we found that neither MS334 nor In9*pvcrt* could predict the clinical outcome following CQ treatment.

The genetic cross conducted by Sá and colleagues identified variable TGAAGH motif lengths at MS334 and In9*pvcrt* as markers of *in vivo* CQ efficacy (in an *Aotus* host) in CQS and CQR progeny, and that CQR progeny had increased expression of *pvcrt*-o (12). However, it is unclear whether either of these indels has a causative role in determining the response to CQ through increased *pvcrt*-o expression or other mechanisms. Interestingly, a study of gene expression across lifecycle stages in *P. vivax* isolates from Cambodia showed that *pvcrt*-o has multiple potential protein isoforms due to the retention of intron 9 that predicted an early stop codon (23). Another study, conducted on clinical isolates from Thailand and Indonesia, identified an AAG trinucleotide insertion in the *pvcrt-o* exon 1 encoding an extra lysine, associated with reduced CQ IC_50_s, although the insertion was observed predominantly in Thai isolates (76%) rather than Indonesian isolates (2.2%); the study was potentially confounded by population structure effects (7). TGAAGH motifs have been shown to regulate gene expression in *Arabidopsis*, but there is currently no knowledge regarding their role in *Plasmodium* (24, 25). In Malaysia, we observed a wide variety of TGAAGH motif lengths in both MS334 and In9*pvcrt* amongst CQS and CQR isolates, with no evidence of a correlation between motif length and clinical outcome following treatment with CQ. For example, the Malaysian CQR isolate TV005 possessed the shortest TGAAGH motif lengths in both gene regions, while one CQR isolate collected at recurrence, MK2079, possessed the longest motif lengths in both gene regions.

Although increased *pvcrt-o* expression has been associated with CQ resistance in several studies (10-12), this pattern has not been replicated in clinical datasets of populations with high-grade CQ resistance. Study of clinical *P. vivax* isolates from Papua, Indonesia, an area of high-grade CQ resistance, found no evidence of association between *pvcrt-o* expression and *ex vivo* CQ susceptibility (14). Furthermore, although a population genomic study using phenotype-free methods to detect selective sweeps identified a weak signal of selection in the vicinity of *pvcrt-o* in an Ethiopian population, this signal was not identified in similar studies conducted in either Malaysia or Papua, Indonesia, where high grade CQ resistance is present (26-28). It is possible that different CQ resistance mechanisms are involved in different *P. vivax* populations. Alternatively, the CQ resistance mechanism may not be heritable, in which case studies applying population genomic approaches to detect sweeps or association methods using pre-treatment clinical isolates may fail to identify a signal.

In the current study, we analysed paired isolates collected at baseline and again on recurrence to explore any changes in the TGAAGH motif lengths after drug exposure. Although the sample size was limited, there was no evidence of an association between MS334 or In9*pvcrt* motif length and CQ resistance. Six paired isolates had different MS334 motif lengths pre and post treatment, however the direction of change (i.e., increase versus decrease in motif length) was inconsistent. Microsatellite-based genotyping data confirmed that the post treatment isolates were the same strains as those present at pre-treatment, except for two pairs, MV019/MV1019 and TK026/TK1026; in these pairs, the isolate at baseline was monoclonal, but the isolates at recurrence comprised at least two strains. It’s plausible that a polyclonal infection was present at both baseline and recurrence, but a minor CQR strain was only detected at recurrence following preferential growth under selective drug pressure. Malaysia is in the pre-elimination stage at the time of the trial, with intense drug pressure resulting in *P. vivax* population bottlenecking, and thus *P. vivax* CQR strains could be present and circulating in the population as the last remaining parasites (26). CQ was the first-line treatment throughout the study period and in view of its very poor efficacy, in 2016 national antimalarial policy was changed to recommend artemisinin combination therapies for the treatment of *P. vivax*.

Our study has several limitations. The extensive sequence complexity in the MS334 and In9*pvcrt* regions, and the high diversity of variants observed in the Malaysian population imposed challenges in sequence alignment and variant calling. To overcome these challenges, we confined our study to high quality monoclonal infections and two independent investigators, both blinded to the sample’s CQ sensitivity status, analysed the molecular assemblies. We were unable to rule out the potential impact of host or other parasitological factors that might impact CQ treatment outcome in the Malaysian study. All patients treated with CQ in our study had observed administration with correct dosage based on body weight and confirmed therapeutic plasma CQ concentrations at day 7 (16). At the time of recurrence, 9 of 20 patients assessed in the original study had plasma CQ + DCQ concentrations in excess of 15 ng/mL (range, 19.7–to 120.5), demonstrating parasite growth in the presence of adequate drug concentrations (16). By definition, these parasites were CQR. Furthermore, the risk of recurrence by day 28 exceeded 60%, far higher than that expected under the conditions of a clinical trial (3). Our study was also limited by small sample size, and despite the near-equal proportions of the two treatment outcome groups, there was no trend in our findings.

In summary, our study found that the MS334 and In9*pvcrt* markers were not effective predictors of clinical outcome following CQ treatment of patients with *P. vivax* malaria in Malaysia. Further studies exploring the mechanism of *P. vivax* CQ resistance and the identification of molecular markers using candidate-free genome-wide approaches are warranted.

## Data Availability

All data produced in the present work are contained in the manuscript

## Acknowledgements

We thank the participants in this study; the Malaysian clinical and laboratory research staff; the Director-General of Health, Malaysia; Kim Piera and Ammar Aziz for conducting the polymerase chain reaction for species diagnosis; and Irene Handayuni for conducting the microsatellite genotyping. The study was funded by the National Health and Medical Research Council (NHMRC) Australia Ideas Grant (APP2001083) awarded to SA, program Grant 1037304, project grant 1045156, and a Senior Principal Research Fellowship to NMA (1135820)], Malaysian Ministry of Health (grant BP00500420), and the AusAID Asia-Pacific Malaria Elimination Network (grant 108-07). The work was also supported by the Australian Centre for Research Excellence on Malaria Elimination (ACREME), funded by the National Health and Medical Research Council of Australia (APP1134989).

---

**Supplementary File 2.**
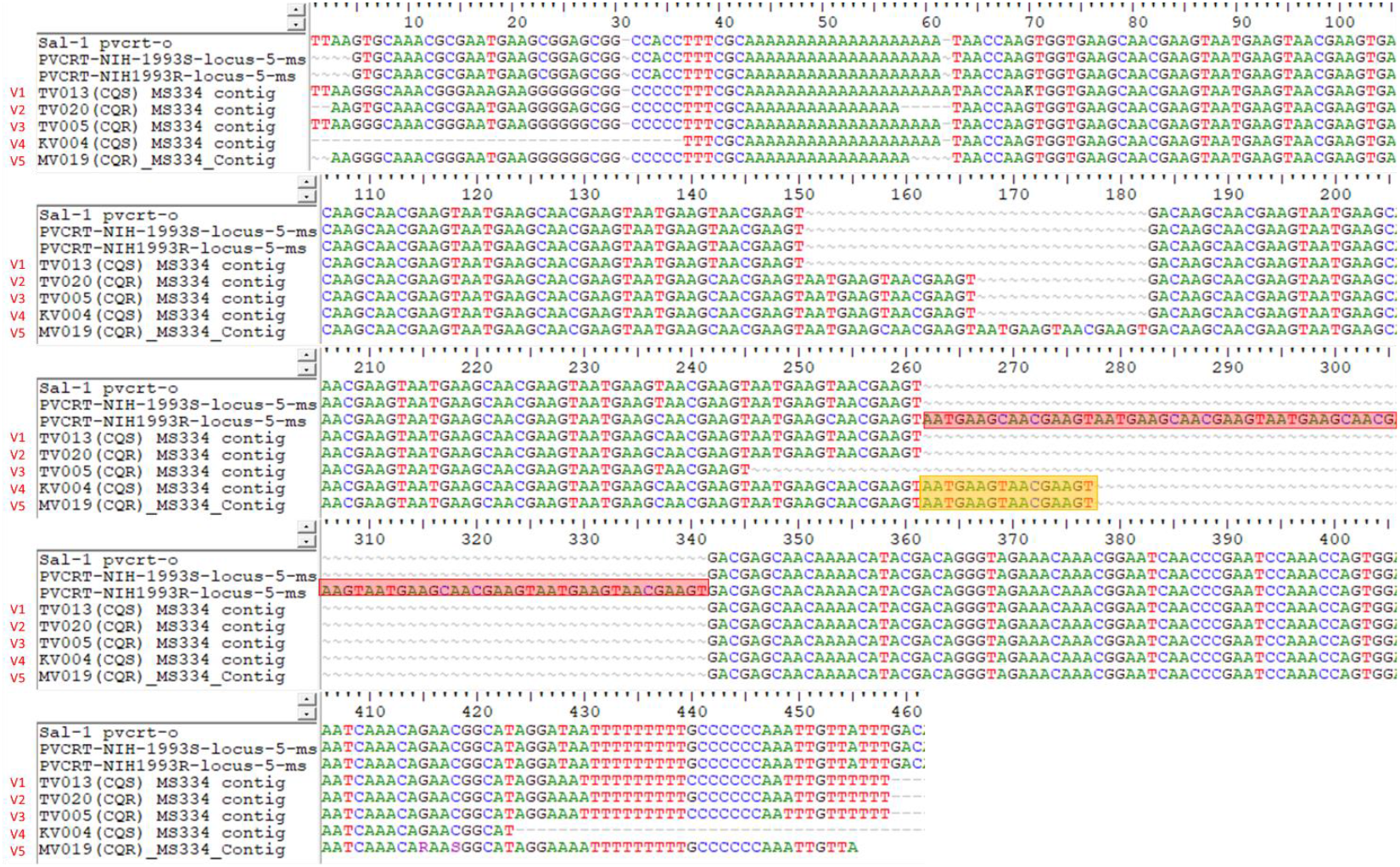
Aligned sequences from the flanking region of *pvcrt-o* in *P. vivax* Sal-1 strain, NIH-1993-S, NIH-1993-R and 5 variants of Malaysian MS334 amplicons. Positions shown are 200bp upstream from the start of *pvcrt-o* in the Sal-1 strain (Pv_Sal1_chr01:330,260). Insertion observed in chloroquine resistant progeny NIH-1993-R is highlighted in red, while region of Malaysian amplicons that contain part of the insertion is highlighted in yellow. V1 to V5 refers to the MS334 allele variant number observed in Malaysian population (see **Table 3**).

**Supplementary File 3.**
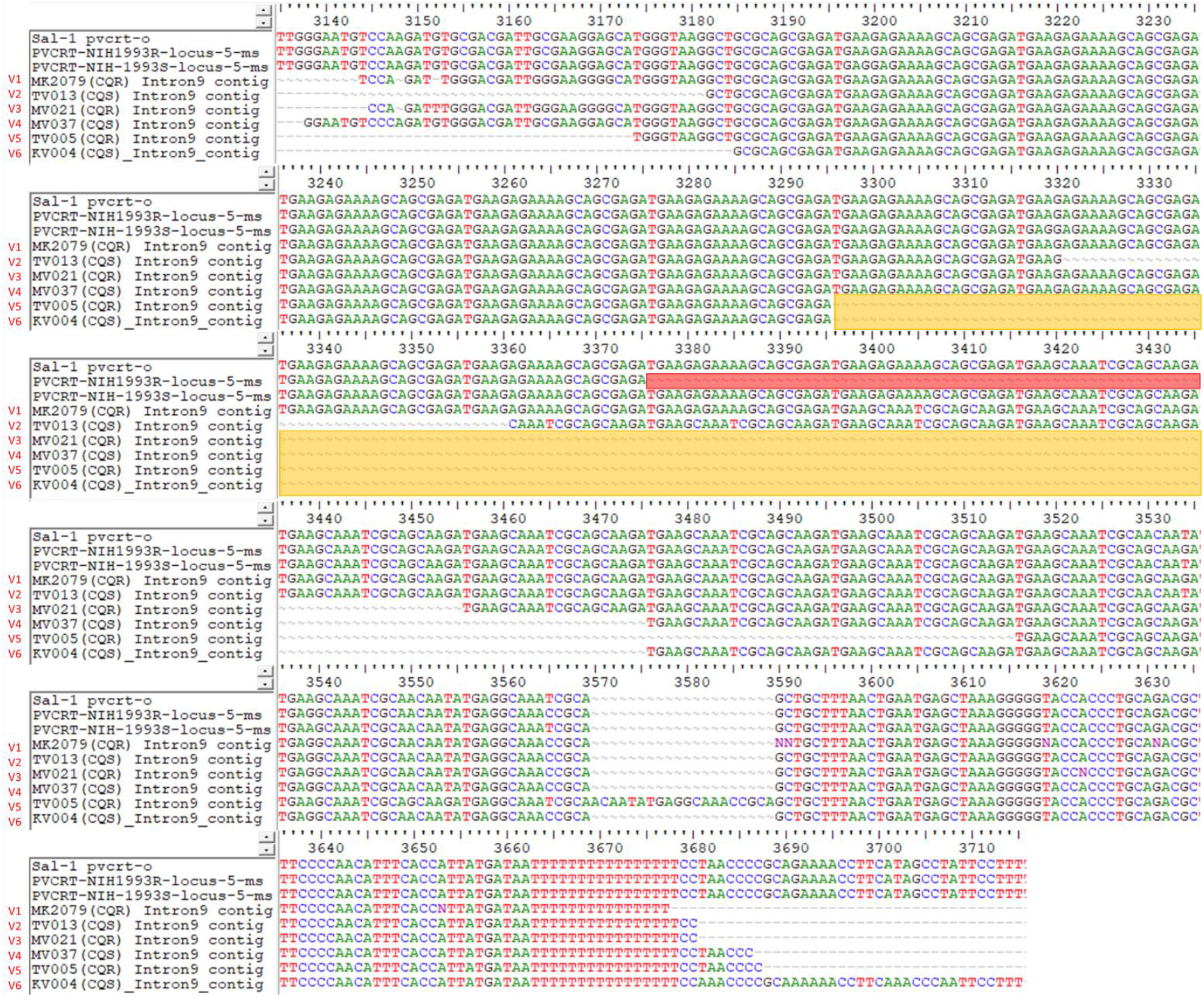
Aligned sequences of *pvcrt-o* Intron 9 region of *P. vivax* Sal-1 strain, NIH-1993-S, NIH-1993-R and 6 variants of Malaysian isolates. Positions shown are approximately 2.8kbp downstream from the start of *pvcrt-o* in Sal-1 strain (Pv_Sal1_chr01:330,260). Deletion observed in chloroquine resistant progeny NIH-1993-R is highlighted in red, while region of Malaysian amplicons that contain part of the deletion is highlighted in yellow. V1 to V6 refers to the *In9pvcrt* allele variant number observed in Malaysian population (see **Table 3**).

